# Impact of Ambient Artificial Intelligence Notes on Provider Burnout

**DOI:** 10.1101/2024.07.18.24310656

**Authors:** Jason Misurac, Lindsey A. Knake, James M. Blum

## Abstract

**Background:** Healthcare provider burnout is a critical issue with significant implications for individual well-being, patient care, and healthcare system efficiency. Addressing burnout is essential for improving both provider well-being and the quality of patient care. Ambient artificial intelligence (AI) offers a novel approach to mitigating burnout by reducing the documentation burden through advanced speech recognition and natural language processing technologies that summarize the patient encounter into a clinical note to be reviewed by clinicians.

**Objective:** To assess provider burnout and professional fulfilment associated with Ambient AI technology during a pilot study, assessed using the Stanford Professional Fulfillment Index (PFI).

**Methods:** A pre-post observational study was conducted at University of Iowa Health Care with 38 volunteer physicians and advanced practice providers. Participants used a commercial ambient AI tool, over a 5-week trial in ambulatory environments. The AI tool transcribed patient-clinician conversations and generated preliminary clinical notes for review and entry into the electronic medical record. Burnout and professional fulfillment were assessed using the Stanford PFI at baseline and post-intervention.

**Results:** Pre-test and post-test surveys were completed by 35/38 participants (92% survey completion rate). Results showed a significant reduction in burnout scores, with the median burnout score improving from 4.16 to 3.16 (p=0.005), with validated Stanford PFI cutoff for overall burnout 3.33. Burnout rates decreased from 69% to 43%. There was a notable improvement in interpersonal disengagement scores (3.6 vs. 2.5, p<0.001), although work exhaustion scores did not significantly change. Professional fulfillment showed a modest, non-significant increase (6.1 vs. 6.5, p=0.10).

**Conclusions:** Ambient AI significantly reduces healthcare provider burnout and modestly enhances professional fulfillment. By alleviating documentation burdens, ambient AI improves operational efficiency and provider well-being. These findings suggest that broader implementation of ambient AI could be a strategic intervention to combat burnout in healthcare settings.

## 1. BACKGROUND AND SIGNIFICANCE

The phenomenon of healthcare provider burnout has garnered significant attention due to its widespread prevalence and profound implications on individual healthcare workers, patient care, and overall healthcare system efficiency^1,2^. Burnout among healthcare providers is characterized by a triad of symptoms: emotional exhaustion, depersonalization, and a reduced sense of personal accomplishment ^2^. These symptoms not only affect the mental and physical health of the providers but also compromise patient safety, increase the likelihood of medical errors, and lead to higher staff turnover rates.^1-3^ Thus, addressing provider burnout is not only a matter of improving individual well-being but also enhancing the quality and safety of patient care.^4^

Ambient artificial intelligence (AI) represents an innovative approach to reducing one of the key contributors to burnout: the clinical burden of documentation. Ambient AI systems utilize advanced speech recognition and natural language processing technologies to transcribe the clinician-patient dialogue and transform it into the appropriate clinical documentation format, thereby reducing the time providers spend on documentation tasks.^5-7^ This shift has the potential to allow providers more time for direct patient care and personal engagement, which are often cited as more fulfilling aspects of healthcare professions.^3,8^

## 2. OBJECTIVES

This prospective observational study examines the impact of ambient AI on reducing burnout and enhancing professional fulfillment among healthcare providers, utilizing a standardized burnout inventory as the primary evaluative tool. The tool measures both the positive aspects of professional fulfillment, including personal accomplishment and work-life balance, and the negative aspects, such as burnout and work exhaustion. By implementing ambient AI amongst a diverse provider base and tracking its influence, the study aims to provide evidence of the technology’s effectiveness on provider well-being.

## 3. METHODS

This was a pre-post, observational study assessing the impact of an ambient listening technology on provider burnout and professional fulfillment at University of Iowa Health Care (UIHC). The University of Iowa Institutional Review Board (IRB) reviewed the project and determined that it did not constitute human subjects research as defined under relevant regulations.

Thirty-eight volunteer physicians and advanced practice providers were selected to participate in a trial of a commercial ambient AI tool (Nabla Copilot, Nabla, Paris, France). The participants included a diverse mix of physicians, nurse practitioners, and physician assistants across multiple specialties. They participated in a 5-week trial of the ambient AI tool.

The tool functions by automatically transcribing patient-clinician conversations and utilizing generative artificial intelligence to create preliminary clinical notes. These notes were reviewed by the participants, edited as appropriate, and entered into the electronic medical record. The AI tool was reviewed by the UIHC compliance and security teams, confirming compliance with the Health Insurance Portability and Accountability Act (HIPAA) as well as UIHC security protocols. Of note, electronic health record (EHR) integration was not pursued during the initial evaluation due to local information technology (IT) constraints, but integration of the ambient AI into the EHR is available. To create the note, providers used a mobile communication device and the AI tool’s website to record the conversation and create the note. The note was then copied into the UIHC EHR (Epic Systems, Verona, Wisconsin, USA).

The primary outcome measure was provider burnout and professional fulfilment, assessed using the Stanford Professional Fulfillment Index (PFI).^2^ The PFI is a validated instrument that measures 3 key dimensions: professional fulfillment, work exhaustion, and interpersonal disengagement. These collectively provide a comprehensive view of a healthcare providers professional quality of life. The scores were scaled to a 10-point scale. Using this scale, the cutpoint for professional fulfillment is 7.5 and greater, and for overall burnout is 3.325 and greater.^2^ Those with average scores beyond these cutpoints are more likely to be professionally fulfilled or to be experiencing burnout, respectively.^2^

The overall burnout score of the PFI has two components: work exhaustion and interpersonal disengagement. Questions within the work exhaustion section focus on physical/emotional exhaustion, a lack of energy at work, and a sense of dread about work-related tasks. The interpersonal disengagement section asks respondents to report their level of connection with and empathy towards patients and colleagues.

Burnout scores were collected at baseline (pre-intervention) and after the five-week trial (post-intervention). Surveys were conducted using an electronic survey tool (Qualtrics) and data were collected in an identifiable fashion for further discussion where required.

Secondary outcome measures included a qualitative assessment of the clinical notes generated by Nabla Copilot, along with a feedback survey for participants which captured their experiences, satisfaction, and any challenges encountered when using the tool.

### 3.1. Statistical Analysis

A minimum sample size of 30 was calculated as necessary to detect a 25% reduction in burnout with a power of 80% and significance of p=0.05. Pre- and post-burnout scores were analyzed using the Related-Samples Sign Test. Statistical analyses were carried out using SPSS 29.0, with a significance level set at p < 0.05. Power analysis was conducted with GPower 31.

## 4. RESULTS

### 4.1. Cohort Description

Pre-test and post-test surveys were completed by 35/38 participants for a 92% survey completion rate. Characteristics of the survey respondents are detailed in Table 1 and their medical area of specialty is listed in Table 2. Utilization of the ambient AI assistant varied significantly among users, with a median of 51 patient encounters in the 5-week trial (interquartile range 17-85). Average visit duration was 18 minutes per visit.

**Table 1:**
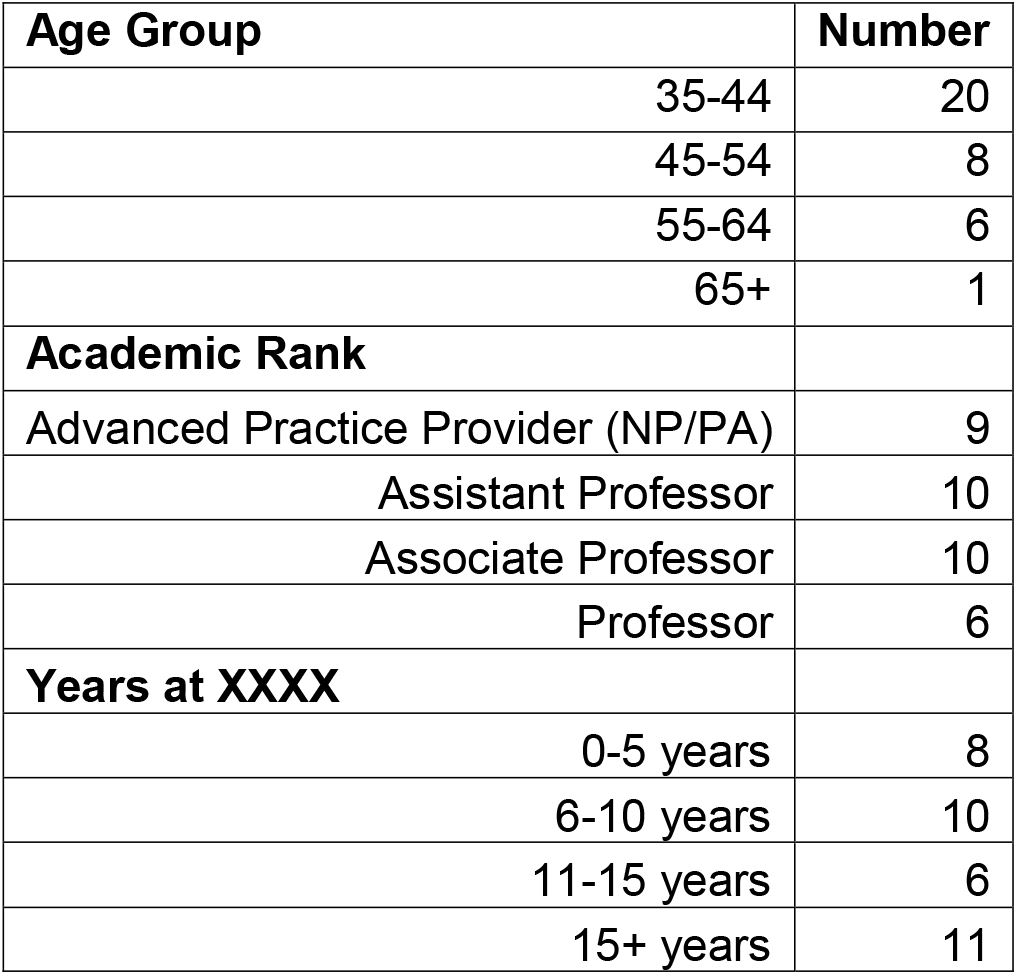
Characteristics of respondents to both pre and post survey (n=35)

**Table 2:** Survey respondents’ subspecialty (n = 35)

| Specialty | Number |
| --- | --- |
| **Family Medicine** | 7 |
| **Internal Medicine** |  |
| Cardiology | 1 |
| Endocrinology | 1 |
| General Internal Medicine | 2 |
| Infectious Disease | 2 |
| Hematology/Oncology | 3 |
| Rheumatology | 2 |
| **Pediatrics** |  |
| Endocrinology | 3 |
| Gastroenterology | 2 |
| Neurology | 1 |
| Pediatric Nephrology | 3 |
| Rheumatology | 1 |
| OB/GYN | 4 |
| PM&R | 1 |
| Head and Neck Oncology | 1 |
| Sports Medicine and Urgent Care | 1 |

OB/GYN (obstetrics and gynecology), PM&R (physical medicine and rehab)

### 4.2. Burnout Analysis

The trial showed a significant reduction in burnout as measured by the Stanford PFI. The median burnout score improved from 4.16 to 3.16 (p=0.005), see Figure 1. The validated cutoff for overall burnout using the PFI is 3.325 on a 10-point scale. Using this cutoff, the burnout rate decreased from 69% (24/35) to 43% (15/35). Given the variation in how often participants used ambient AI, we subdivided the group into quartiles by utilization of ambient AI and calculated the pre- and post-burnout scores in each quartile (Fig 2). The lowest quartile of utilization did not demonstrate improvement in burnout scores, while each of the remaining three quartiles had a lower post-ambient AI burnout score.

**Figure 1:**
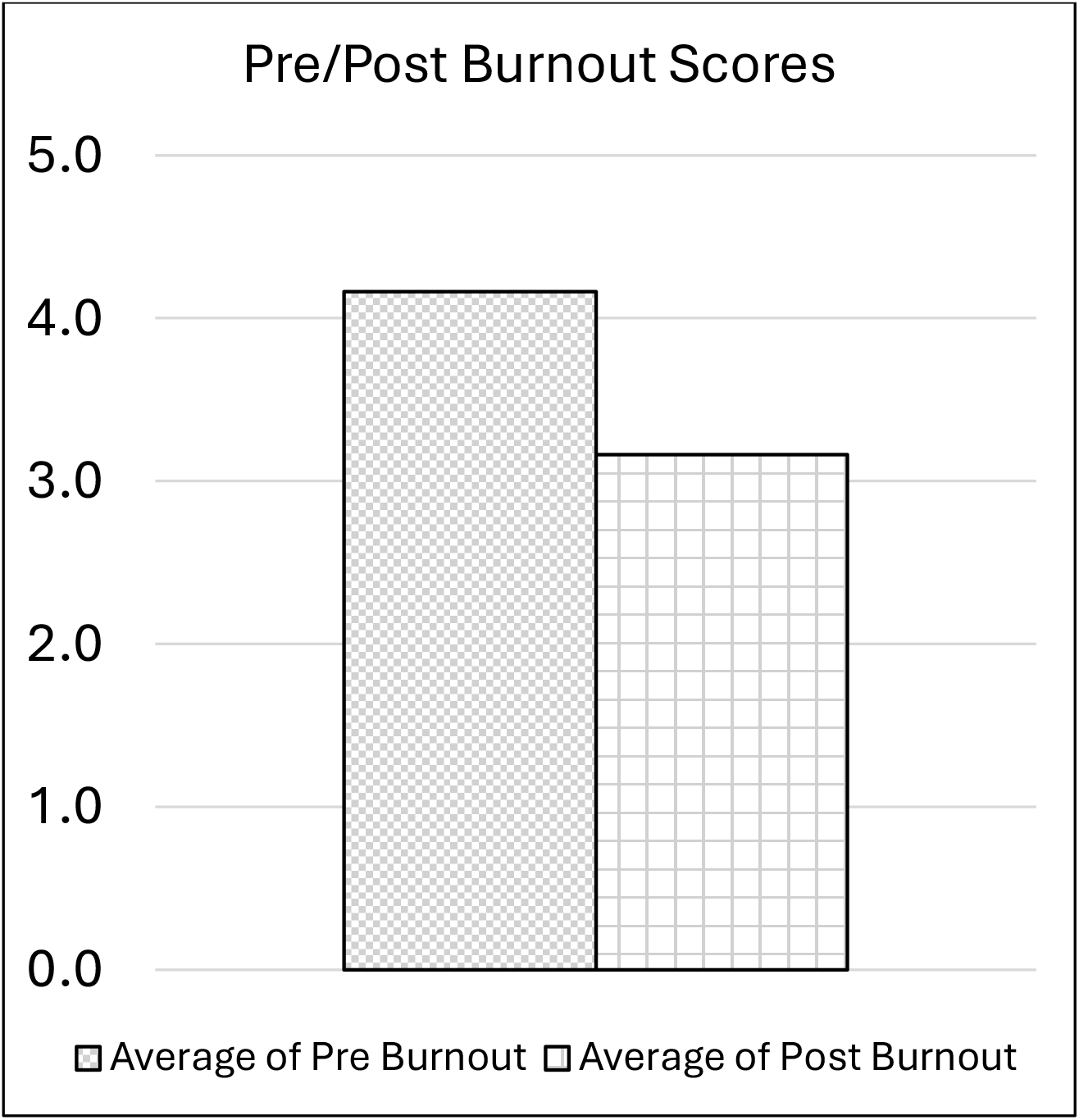
Pre and Post intervention burnout scores, scaled to 10-point scale. Cutoff for burnout using this scale is 3.325.^2^

**Figure 2:**
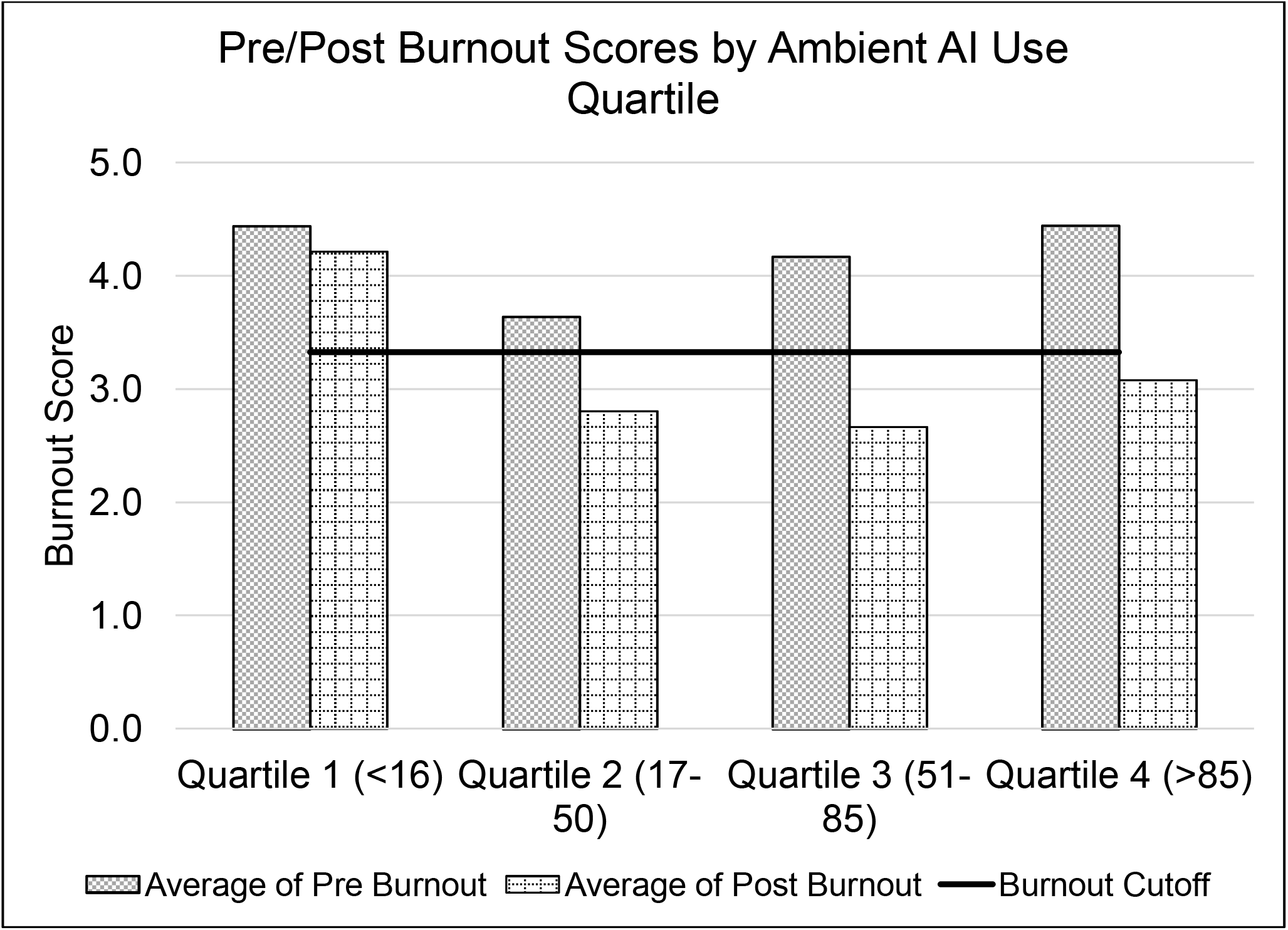
Pre and post intervention burnout scores subdivided by use quartile (number of visits where ambient AI was used), scaled to 10-point scale. Cutoff for burnout using this scale is 3.325. ^2^

We did not observe a statistically significant improvement in work exhaustion scores pre- and post-ambient AI (5.0 vs 4.2, p=0.08). However, there was a robust and significant improvement in interpersonal disengagement scores (3.6 vs 2.5, p<0.001), demonstrating that improvements in interpersonal disengagement were a significant factor in improving overall burnout scores.

Professional fulfillment showed a modest increase in the post-ambient AI survey, though the difference was not statistically significant (6.1 vs 6.5, p=0.10).

### 4.3. User Sentiment and Satisfaction

Twenty-eight out of 35 participants reported a beneficial impact on patient interactions attributed to Nabla Copilot. Specifically, participants highlighted improvements in more focused listening during patient encounters. Additionally, 24/35 reported improvements in their work-life balance, attributing this benefit to modest time savings but more importantly to a reduction in documentation-related stress.

During the trial, users had the opportunity to leave feedback after each note. On average, notes generated by the AI tool were rated 4.3 out of 5.0 stars. Survey comments reflected high satisfaction with the history of present illness (HPI) section, with room for improvement expressed regarding the assessment and plan.

Survey respondents indicated that they appreciated the tool’s ease of use, accurate documentation, customization options, and voice recognition capabilities. However, challenges identified included the lack of seamless EHR integration, the need for yet more personalized customization, and accuracy issues with speaker differentiation. When asked about their level of disappointment if they were unable to use Nabla Copilot in the future, survey respondents had an average score of 7 out of 10, reflecting a strong preference to continue using the tool.

## 5. DISCUSSION

The findings of this trial suggest that ambient AI holds promise as a tool to mitigate physician burnout and enhance patient experiences. This study’s utilization of the Stanford PFI revealed significantly decreased burnout scores and a modest increase in professional fulfillment. The PFI, renowned for its reliability and validity in assessing both the positive aspects of professional fulfillment and the negative aspects of burnout, provided a comprehensive framework for this analysis. Among the 35 healthcare providers participating, those who used ambient AI for at least 17 patient encounters showed a substantial improvement in their PFI burnout scores.

### 5.1. Reduction in Burnout Symptoms

The decrease in burnout as measured by the PFI can be attributed to several mechanisms facilitated by ambient AI. Firstly, ambient AI has been demonstrated to significantly reduce the time providers spend on documentation, which is directly correlated with lower burnout rates.^9,10^ Secondly, ambient AI’s efficiency likely allows providers to engage more meaningfully with their patients, improving interpersonal engagement and enhancing job satisfaction—a key component of professional fulfillment. These results align with prior research indicating that reductions in non-clinical administrative tasks lead to higher levels of professional fulfillment and lower burnout.^11,12^

The qualitative feedback provided by participants offers valuable insights into the strengths and limitations of ambient AI tools from the end-user perspective. While many appreciated the tool’s ease of use and ability to capture visit details, others expressed concerns regarding the need for manual editing to align the output with their preferred documentation style. This feedback provides some guidance to iterative development of ambient AI, which should be sufficiently customizable to provide the level of detail and documentation style for a wide variety of specialties and outpatient settings. This clinician feedback is similar to what has been published in other types of AI-technology used in healthcare, such as AI-drafted patient messages responses where PFI burnout scores were reduced but clinician time was not significantly reduced due to need for editing/optimization.^13,14^ Simulated physician and patient interactions have been given to large language models like ChatGPT-4 to create clinical notes and discharge summaries and have been found to have a substantial number of omission errors, hallucinations, and poor note quality.^15^ Thus, AI technologies must continue to evolve and adapt to the different clinical use cases and needs. However, overall, this study supports the notion that existing ambient AI tools will positively impact provider burnout metrics if the tools are used frequently, and physicians learn how to adapt to the technology available.

### 5.2. Implications for Clinical Practice

The findings suggest that the integration of ambient AI into healthcare practices could be a strategic intervention to combat provider burnout and enhance professional fulfillment by decreasing the cognitive load required during each patient visit. Healthcare systems should consider broader implementation of ambient AI not only to streamline documentation processes but also to support the well-being of providers. The positive correlation between greater use of ambient AI and improved PFI scores underscores the importance of consistent and sustained use of such technologies to realize their full benefits. Furthermore, the lack of burnout score improvement among users in the lowest quartile of ambient note usage provides additional evidence that the improvement in participant burnout scores can be attributed to use of the ambient AI tool.

The data from this study also supports the notion that comprehensive integration of the tool into the EHR is not required to potentially impact provider burnout. The use of the ambient AI tool required clinicians to work with a mobile device and separate website to use the technology. Despite this burden, a positive impact was seen in the PFI.

### 5.3. Limitations and Future Research

Despite promising results, this study has several limitations. This study was a single-center, observational, pre-post pilot study over a short period of time, which was powered to demonstrate a difference in PFI, and while a difference was demonstrated, there is still the possibility of type I error. In addition, the providers included in this evaluation volunteered for this study, thus, they may not be representative of providers in general and do not cover the breadth of all clinical subspecialties. Future research should aim to include a larger cohort and possibly utilize a randomized controlled design to validate these findings across varied healthcare settings and populations. This study did not incorporate a formal assessment of time spent in notes, but future work will include a formal time in notes analysis once this application is integrated into the EHR. It is also essential to conduct longitudinal studies to assess the long-term impacts of ambient AI on professional fulfillment and burnout.

Exploring the relationship between ambient AI usage and other factors such as patient outcomes and clinical efficiency would provide a more holistic understanding of its benefits and limitations. Future work should include incorporating patient survey data to understand patient opinions and fears on incorporating this new technology. However, preliminary research reports that both patients and physicians find ambient technology improves their healthcare experiences^10^. Additionally, further studies could investigate the integration process of such technologies and the potential barriers to their adoption in different healthcare environments as well as a cost-benefit analysis.

Future work should focus on the significance of exploring how ambient AI’s potential extends beyond individual benefits to broader systemic implications. In a healthcare environment increasingly pressured by high demands and limited resources, the ability of technology to mitigate burnout can lead to better health outcomes, reduced costs associated with staff turnover and absenteeism, and a more sustainable work environment for providers.

## 6. CONCLUSIONS

In conclusion, this observational study indicates that ambient listening technology can play a critical role in reducing provider burnout as quantified by the Stanford PFI. By alleviating the burden of documentation and cognitive load required by the clinicians during a patient visit, ambient AI not only improves operational efficiencies but also enhances the work-life balance of healthcare providers. This innovation holds the potential to significantly impact the future dynamics of healthcare delivery and provider satisfaction.

## Data Availability

All data produced in the present study are available upon reasonable request to the authors

## ACKNOWLEDGEMENTS

None.

## REFERENCES

1. Shanafelt TD, West CP, Dyrbye LN, et al. Changes in Burnout and Satisfaction With Work-Life Integration in Physicians During the First 2 Years of the COVID-19 Pandemic. Mayo Clin Proc. Dec 2022;97(12):2248–2258. doi:10.1016/j.mayocp.2022.09.002

2. Trockel M, Bohman B, Lesure E, et al. A Brief Instrument to Assess Both Burnout and Professional Fulfillment in Physicians: Reliability and Validity, Including Correlation with Self-Reported Medical Errors, in a Sample of Resident and Practicing Physicians. Acad Psychiatry. Feb 2018;42(1):11–24. doi:10.1007/s40596-017-0849-3

3. West CP, Dyrbye LN, Shanafelt TD. Physician burnout: contributors, consequences and solutions. J Intern Med. Jun 2018;283(6):516–529. doi:10.1111/joim.12752

4. Shanafelt TD, Schein E, Minor LB, Trockel M, Schein P, Kirch D. Healing the Professional Culture of Medicine. Mayo Clin Proc. Aug 2019;94(8):1556–1566. doi:10.1016/j.mayocp.2019.03.026

5. Ghatnekar S, Faletsky A, Nambudiri VE. Digital scribe utility and barriers to implementation in clinical practice: a scoping review. Health Technol (Berl). 2021;11(4):803–809. doi:10.1007/s12553-021-00568-0

6. Crampton NH. Ambient virtual scribes: Mutuo Health’s AutoScribe as a case study of artificial intelligence-based technology. Healthc Manage Forum. Jan 2020;33(1):34–38. doi:10.1177/0840470419872775

7. Coiera E, Kocaballi B, Halamka J, Laranjo L. The digital scribe. NPJ Digit Med. 2018;1:58. doi:10.1038/s41746-018-0066-9

8. Friedberg MW, Chen PG, Van Busum KR, et al. Factors Affecting Physician Professional Satisfaction and Their Implications for Patient Care, Health Systems, and Health Policy. Rand Health Q. Winter 2014;3(4):1

9. Balloch J, Sridharan S, Oldham G, et al. Use of an Ambient Artificial Intelligence Tool to Improve Quality of Clinical Documentation. FHJ. 2024;In pressdoi:10.1016/j.fhj.2024.100157

10. Tierney AA, Gayre G, Hoberman B, et al. Ambient Artificial Intelligence Scribes to Alleviate the Burden of Clinical Documentation. NEJM Catal Innov Care Deliv. 2024;5(3)doi:10.1056/CAT.23.0404

11. Erickson SM, Rockwern B, Koltov M, McLean RM, Medical P, Quality Committee of the American College of P. Putting Patients First by Reducing Administrative Tasks in Health Care: A Position Paper of the American College of Physicians. Ann Intern Med. May 2 2017;166(9):659–661. doi:10.7326/M16-2697

12. Swensen S, Shanafelt T. Mayo Clinic Strategies To Reduce Burnout: 12 Actions to Create the Ideal Workplace. Oxford University Press; 2020. 10.1093/med/9780190848965.001.0001

13. Garcia P, Ma SP, Shah S, et al. Artificial Intelligence-Generated Draft Replies to Patient Inbox Messages. JAMA Netw Open. Mar 4 2024;7(3):e243201. doi:10.1001/jamanetworkopen.2024.3201

14. Tai-Seale M, Baxter SL, Vaida F, et al. AI-Generated Draft Replies Integrated Into Health Records and Physicians’ Electronic Communication. JAMA Netw Open. Apr 1 2024;7(4):e246565. doi:10.1001/jamanetworkopen.2024.6565

15. Kernberg A, Gold JA, Mohan V. Using ChatGPT-4 to Create Structured Medical Notes From Audio Recordings of Physician-Patient Encounters: Comparative Study. J Med Internet Res. Apr 22 2024;26:e54419. doi:10.2196/54419

